# Evaluating the Impact, Implementation and Sustainability of the Suicide Prevention Grant Fund – A Qualitative Study

**DOI:** 10.64898/2025.12.22.25342818

**Authors:** Dominika Valacsay, Jordan Thompson, Kirsten Anstey, Rina Dutta, Kathleen Fraser, Andrew Grundy, Winnie Liu, Patrick Nyikavaranda, Karen Machin, Alan Simpson, Sonia Johnson, Rebecca Appleton, Brynmor Lloyd-Evans

## Abstract

**Background:** Short-term funding initiatives are often used to stimulate innovation and improve service delivery. They are a main source of funding for voluntary, community and social enterprise (VCSE) organisations. However, reliance on these short-term grants can limit their funded activities and sustainability. In 2023, the Department of Health and Social Care (DHSC) set up the Suicide Prevention Grant Fund (SPGF). This was awarded to VCSEs for a single year to deliver diverse, innovative interventions aimed at preventing suicide. This study aimed to explore service managers’ perceptions of the SPGF’s impact. Specifically, we sought to identify key factors that contributed to a successful delivery of the projects, any challenges in the implementation process and the services’ capacity to carry on beyond the funding period.

**Methods:** We conducted semi-structured online interviews with managers from 20 SPGF-funded services across England. Representatives from an additional 31 services completed a short survey to complement the interview data. Data were analysed using a framework analysis, with a combination of inductive and deductive coding. The analytic framework was developed and refined in collaboration with our expert working group, including our team of Lived Experience Researchers.

**Results:** Data were categorised into three overarching themes, shaped by our objectives (impact, implementation, and sustainability), with 16 sub-themes identified across these themes. Funding had a clear impact in expanding service provision. Managers identified the importance of the funded projects, which often filled gaps in service provision, targeting underserved groups. Many reported improved well-being, coping, and a reduction in distress. Implementation facilitators included co-designed interventions, skilled staffing, and productive partnerships with external agencies. However, short timescales, administrative burden, and a retrospective payment model particularly strained smaller charities. Sustainability outlooks were mixed. A minority (notably digital or embedded services) expected full continuation; most anticipated partial continuation or closure without further funding.

**Conclusions:** Short-term grants can catalyse impactful, targeted suicide prevention, but sustainability is jeopardised by brief funding windows, reimbursement models, and administrative load, especially for smaller VCSEs. Many services indicated perceived benefits of their interventions, but further evaluation of the impact of novel initiatives in reducing suicide rates is desirable, where possible.

## Background

Suicide remains a significant public health concern globally, ranking among the leading causes of preventable death [1]. Sadly, suicide is still the third leading cause of death worldwide for individuals aged 15-29 [2].

According to the Office for National Statistics [3], the suicide rate in England and Wales increased by 7.57% from 2022 to 2023 and was the highest rate since 1999 for both men and women. The most recent report measuring suspected suicide rates for January 2025 [4] showed 11.0 deaths per 100,000 people. The primary individual risk factors include a history of self-harm or suicide attempts, being male and middle-aged, and having a psychiatric or chronic physical illness, particularly those involving pain [5,6] In 2023, 74% of the 6,069 registered suicides in the UK were among middle-aged men, with the highest risk observed in those aged 45–64, making this group the most at risk of suicide [7]. Social factors like divorce, living alone, and unemployment also increase risk, as do alcohol and drug use . Exposure to another person’s suicide, genetic vulnerability, and specific high-risk occupations, such as construction, entertainment, and caregiving roles in the UK, have also been linked to elevated suicide rates [5, 6].

Suicide prevention strategies target two main areas: preventing psychiatric conditions and addressing specific risk factors like media influence, help-seeking, access to means, and managing self-harm [8]. Restricting access to common means (e.g., medication, pesticides, high-risk locations) has consistently been shown to reduce suicides [9, 10]. One of the biggest challenges in psychiatric suicide prevention is accurate risk assessment, as evidence shows current methods often fall short [11, 12]. According to the National Confidential Inquiry into Suicide and Safety in Mental Health (2021) [13], about 25–30% of people who died by suicide in the UK had contact with mental health services in the year before their death. Half of this group had that final contact within just a week of dying. Nevertheless, 85% were judged by their clinician to be at low or no immediate risk of harm.

The numbers above illustrate two problems. First, most of those who take their lives by suicide are not in touch with mental health services at all, and second, it’s hard to identify those at the highest risk of suicide, even among those who are in contact with mental health services.

They also help to explain why funding voluntary and community organisations is so important, as there is growing support for the idea that VCSEs are often better placed to provide local, accessible, and tailored support that meets people where they are - particularly for marginalised groups that statutory services may struggle to reach [14].

Recognising this, in 2023 the UK government [15] published a suicide prevention strategy called “Suicide prevention in England: 5-year cross-sector strategy”. It emphasised a cross-government approach involving the NHS, local authorities, and community sectors. Goals include reducing the suicide rate within 5 years, with initial declines in half that time, and improving support for self-harm and bereaved individuals. This strategy targets groups most at risk of suicide, as well as aiming to address specific risk factors, including social isolation, gambling and substance misuse.

To achieve these aims, the UK government aims to involve Voluntary, Community, and Social Enterprise organisations (VCSE) in suicide prevention. VCSEs deliver specialised and tailored support at both local and national levels. These organisations are particularly well-placed to provide targeted assistance to the seven high-risk groups identified in the national suicide prevention strategy. In the 2023 Spring Budget, the government announced a £10 million Suicide Prevention Grant Fund (SPGF) [16] to support non-profit organisations in enhancing suicide prevention services amid increasing demand and to fund diverse, innovative initiatives to prevent suicides at both national and community levels. 79 organisations were awarded funding for diverse initiatives, including peer support groups, bereavement counselling, suicide awareness training, community outreach, and digital wellbeing platforms. These initiatives began between December 2023 and January 2024, with all projects completed by 31 March 2025. Short-term funding is a common mechanism for stimulating innovation and expanding a service, particularly in the voluntary and community sector. However, there is a growing idea that these models can limit sustainability, making longer-term funding more desirable. [17].

As one of the largest recent investments in voluntary and community sector suicide prevention in England, the SPGF represents a significant opportunity to understand how government funding can support innovative, locally tailored interventions and what challenges organisations face in sustaining them.

The aim of this study is therefore to explore managers’ perceptions of the projects’ impact, reflections on the implementation process, and views on the projects’ sustainability following the end of the funding. Additionally, the study sought to identify key factors that helped or hindered these processes. The research was commissioned by the DHSC through the NIHR Policy Research Unit in Mental Health (MHPRU).

## Methods

### Ethical considerations

This project was categorised as a service evaluation; therefore formal ethics approval was not required. This was confirmed by the Research and Development team of the North London Research Consortium, after reviewing our study protocol.

### Design

This study employed a qualitative design, utilising semi-structured online interviews, and short form online survey responses with service managers.

### Participants

Since the grant agreements were awarded, one organisation withdrew; in total, 78 organisations received funding from the SPGF initiative. Any service manager, clinical lead or other senior professional involved in delivering or overseeing these initiatives was eligible to take part.

## Interview Methodology

### Sampling

We aimed to gather a diverse and balanced sample that reflected the variety of funded services. Both larger and smaller services were equally valued, and steps were taken to ensure representation across different project types and funding levels.

We purposively selected 30 services to contact for the interview study, to produce a varied sample with respect to:

- **Target population**: i) whole local population; and ii) Suicide Prevention Strategy priority group (Middle-aged men, Perinatal women, Children and Young People); and iii) other social, clinical demographic group (e.g. people already in contact with mental health services, people with previous self-harm experience, people in touch with criminal justice system; autistic people)
- **Provider type**: i) mental health service; and ii) other community servicelJ
- **Bid type**: include at least one Consortium bidlJ(a funding application made collaboratively by two or more organisations)
- **Area** – suicide rate: i) High suicide areas (North West, North East, Yorkshire); ii) lower suicide areas (London) iii) other areas
- **Area – English region**lJ(i.e., the nine standard regions used by the Office for National Statistics: North East, North West, Yorkshire and the Humber, East Midlands, West Midlands, East of England, London, South East, and South West) [3]
- **Service type**: i) clinical or educational intervention (e.g. cancer counselling services, educational toolkits and web platforms, long-term trauma therapy); and ii) social intervention (e.g. peer support groups, community hubs and safe spaces, search and rescue)
- **Service development type**: i) new services; and ii) expansions to existing services
- **Funding amount**: i) Small <£10k; medium £10-100k; large >£100k

We anticipated that not all managers would be willing or able to take part. Recruitment was planned to continue until we reached our target of 20 participants. Our sample size was pre-defined and agreed with the DHSC. The interviews captured a wide range of perspectives across diverse service types, locations, and delivery models, which provided sufficient depth to meet the aims of this service evaluation.

The characteristics of target services were plotted on a grid (see Appendix 1) to allow review of the extent of variation in the planned sampling frame, and to show how the final sample reflected our sampling criteria.

### Data Collection

The topic guide was developed in collaboration with members of the wider working group, which includes topic experts, clinicians, and individuals with personal experience of mental health issues, or of caring for someone with a mental health condition. We also engaged with policymakers from the DHSC SPGF team, who commissioned the projects, to ensure the evaluation was informed by policy priorities. In the topic guide, we asked background questions about the participants’ experiences, and the aims and rationale behind the project. The remainder of the interview guide explored the implementation process, perceived impacts at different levels (service, individual, community), challenges encountered during delivery, adaptations made, and views on sustainability following the end of funding. The full topic guide is provided in Appendix 2.

### Procedures

Potential participants were first informed about the study by the DHSC and made aware that the interviewer was affiliated with the NIHR MHPRU. They were also told that the purpose of the study was to support the DHSC’s evaluation of the SPGF.

Following the services’ interest in participating, the research team contacted them via email with additional information about the study. Once a date and time for the interview had been agreed upon, participants were asked to complete and return a signed consent form prior to the interview. The interviews were conducted via Microsoft Teams, lasting approximately 45–60 minutes, and were video-recorded with the participants’ written consent. Transcripts were generated using Teams’ built-in transcription tool and subsequently checked and corrected by the lead researcher (DV). All identifying information was anonymised during this process. Once transcripts were verified and anonymised, the original video files were permanently deleted.

All interviews were carried out by DV, a postgraduate MSc student and member of the research team, who is trained in qualitative interviewing methods. While most interviews were conducted one-on-one, in three cases, two members of the management team participated. This approach was chosen when participants held responsibility for different components of the funded project, and including two staff members was considered helpful to gather a comprehensive account of how the project was implemented.

### Analysis

We used a mixed deductive–inductive, question-led approach to analysis. The coding was structured around three overarching evaluation questions: (1) what the perceived impacts of the projects were; (2) what helped and hindered project implementation; and (3) how sustainable the projects were after the end of funding, and what factors supported or constrained sustainability.

For implementation (Q2), we developed five main domains deductively from the Consolidated Framework for Implementation Research (CFIR) [18]. The CFIR is a comprehensive framework, widely used in implementation science, with the aim to identify barriers and facilitators to implementation. These domains were: *Intervention characteristics* (e.g., strength, distinctiveness, adaptability), *Outer setting* (e.g., local environment, external partnerships), *Inner setting* (e.g., resources, organisational capacity), *Characteristics of individuals* (e.g., staff skills, commitment), and *Implementation process* (e.g., set-up, delivery, evaluation, adaptation). Within each of these domains, sub-themes were generated inductively from the data using a codebook approach [19] informed by thematic analysis [20. For impact (Q1) and sustainability (Q3), themes and sub-themes were developed inductively, again following a codebook approach. Impact themes focused on how projects were distinctive, responsive to stakeholder needs, and their perceived outcomes. Sustainability themes captured outlooks for continuation, perceived barriers and facilitators, and ongoing support needs.

Coding was initially conducted by the lead researcher (DV) using NVivo (QSR International Pty Ltd, 2020, Version 15) and Microsoft Excel. Four transcripts were reviewed with colleagues from the NIHR MHPRU and lived experience researchers (LERs), who also proposed additional codes based on their reflections. The coding framework was refined iteratively and shared with the LERs for feedback before being applied across the full dataset.

Not all themes were represented across all interviews, as the semi-structured nature of the interviews allowed for varied emphasis based on participants’ roles and experiences.

## Survey Methodology

### Sampling & Data Collection

A survey containing nine questions (plus respondent profile items) was distributed to managers of the 48 SPGF projects not invited to interviews (see Appendix 3). At the start of the survey, managers were presented with some information about the study and how the data will be used; they were required to tick a box to confirm they had read and understood this information before completing the rest of the survey questions. The survey explored managers’ views of their SPGF projects: the extent to which aims were achieved, barriers and facilitators to implementation, and post-grant sustainability.

### Analysis

Open-ended responses were coded deductively using the same framework as the interview analysis described above. Descriptive statistics were compiled to assess how many projects were set to continue after funding ended. We integrated findings from interviews and survey responses in developing and reporting theme summaries.

### Researcher Positionality

The lead researcher (DV) is a postgraduate MSc student in the Clinical Mental Health Sciences programme at University College London (Division of Psychiatry). As part of this dissertation project, DV was responsible for conducting the interviews, reviewing the transcriptions, coding, and conducting the initial analysis.

The research was supervised by two senior researchers (RA, BLE) employed at the NIHR MHPRU, with disciplinary backgrounds in psychology and social work. Two additional researchers (JT, KA) at the NIHR MHPRU completed the analysis of survey responses. In addition, a working group contributed to the project, including a Professor at King’s College London with expertise in suicide research, and three members with lived experience of mental health difficulties and suicide. These three lived experience researchers were actively involved actively involved throughout the study, including in developing the topic guide, reviewing transcripts, and discussions around coding decisions and emergent themes.

The project was commissioned by the DHSC, which requested the evaluation of the SPGF. DHSC staff were involved in developing the interview topic guide to ensure alignment with policy priorities, but had no role in conducting the interviews, analysing the data, or interpreting the findings.

## Results

### Participants

The projects participating in the SPGF employed a variety of prevention strategies, including direct interventions at high-risk locations, provision of mental health support, suicide-specific therapeutic approaches, and community-based initiatives aimed at reducing isolation, such as creating safe spaces for social connection and therefore reducing suicidality indirectly. More details of the interview participants characteristics and the purpose and design of included projects are shown below in Table 1.

**Table 1.**
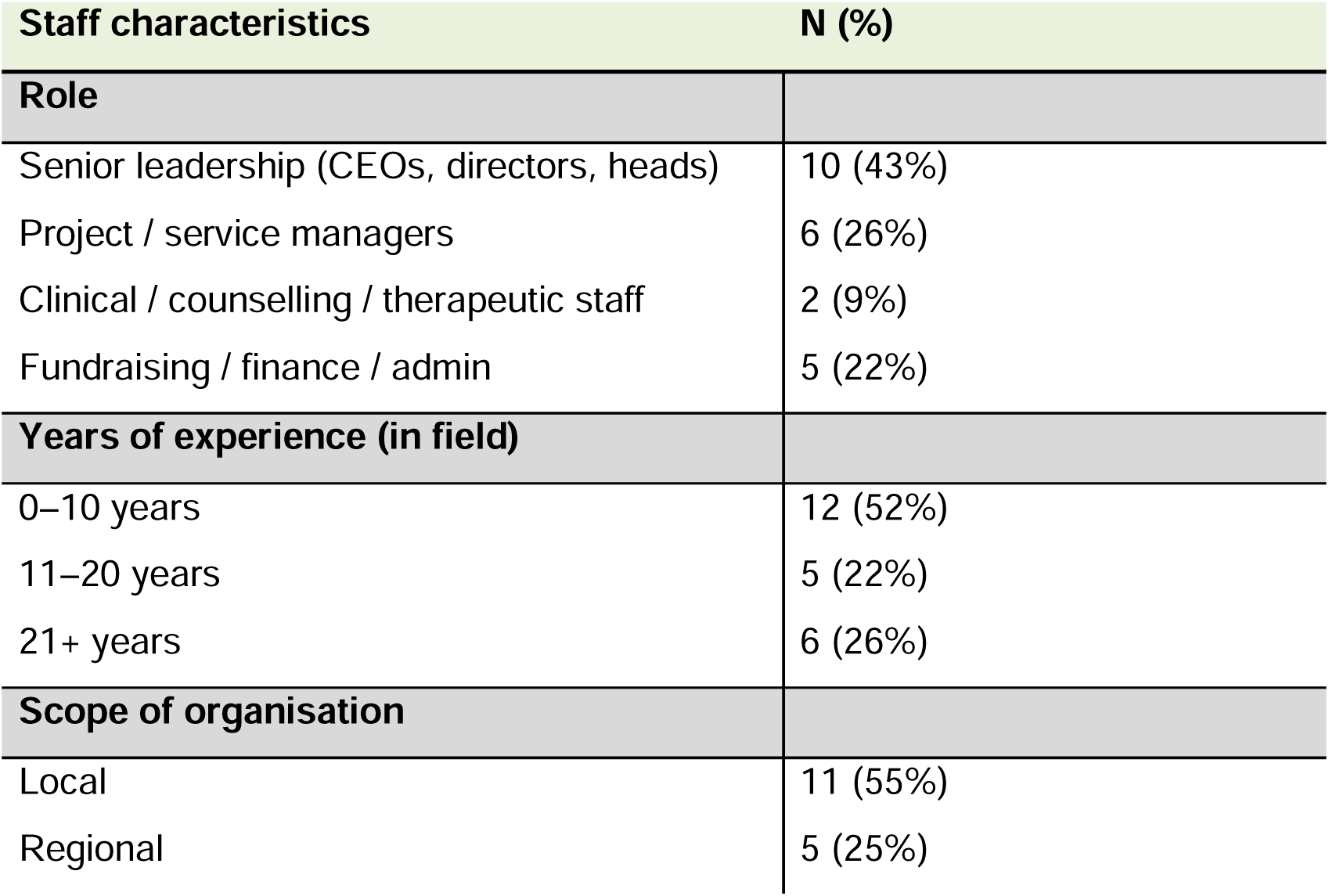
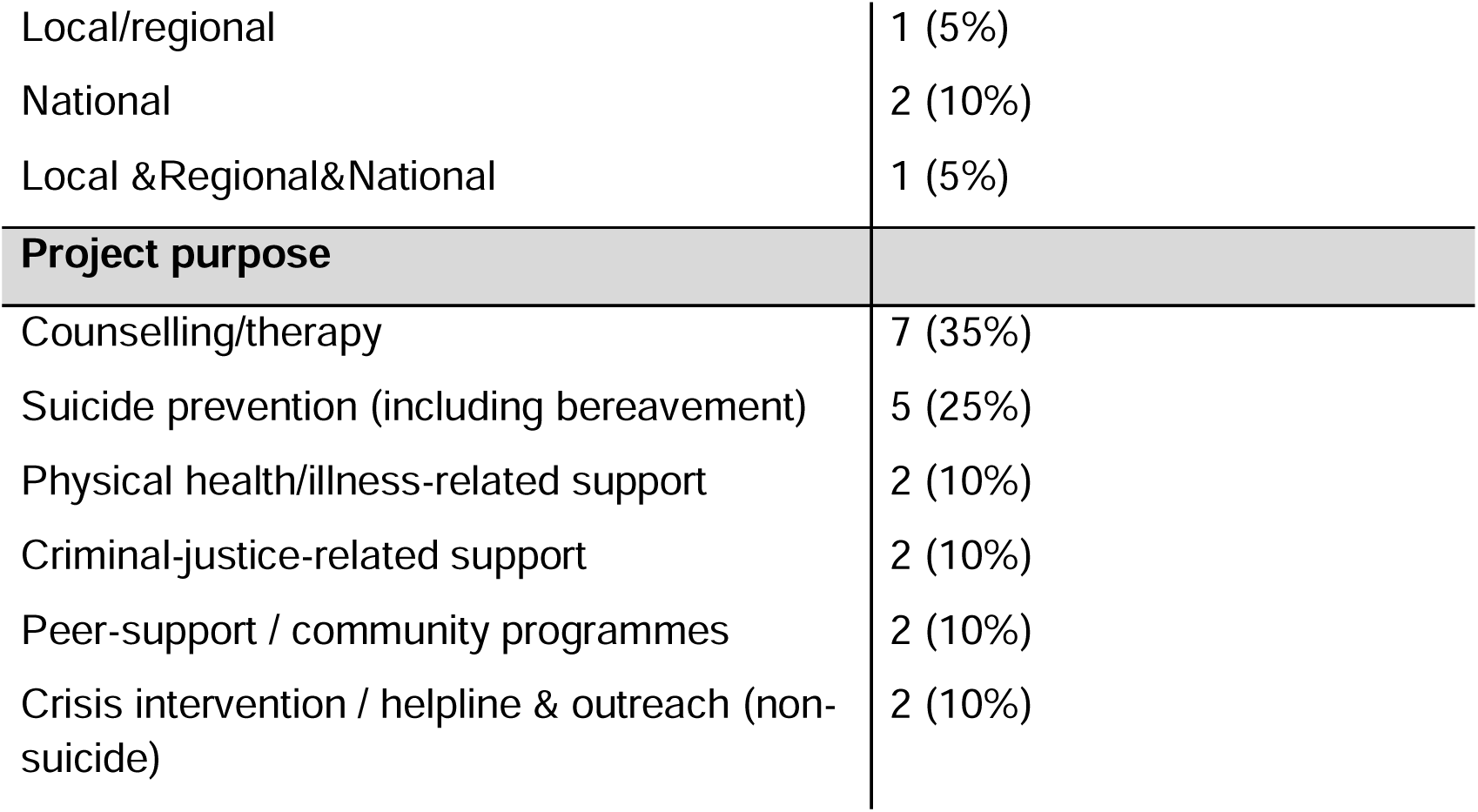
Interview Participant and Project Characteristics. .

| Staff characteristics |  | N (%) |
| --- | --- | --- |
| Role |  |  |
| Senior leadership (CEOs, directors, heads) |  | 10 (43%) |
| Project / service managers |  | 6 (26%) |
| Clinical / counselling / therapeutic staff |  | 2 (9%) |
| Fundraising / finance / admin |  | 5 (22%) |
| Years of experience (in field) |  |  |
| 0–10 years |  | 12 (52%) |
| 11–20 years |  | 5 (22%) |
| 21+ years |  | 6 (26%) |
| Scope of organisation |  |  |
| Local |  | 11 (55%) |
| Regional |  | 5 (25%) |

|  |  |
| --- | --- |
| Local/regional | 1 (5%) |
| National | 2 (10%) |
| Local & Regional & National | 1 (5%) |
| **Project purpose** |  |
| Counselling/therapy | 7 (35%) |
| Suicide prevention (including bereavement) | 5 (25%) |
| Physical health/illness-related support | 2 (10%) |
| Criminal-justice-related support | 2 (10%) |
| Peer-support / community programmes | 2 (10%) |
| Crisis intervention / helpline & outreach (non-suicide) | 2 (10%) |

Survey responses were distributed to all remaining 48 projects not invited to the interview stage. We received 52 responses with 29 of these being complete. Of the incomplete responses, 2 were partially completed and 21 included no analysable data. Duplicate responses from managers were identified in 3 cases, and a further 2 responses had no identifiable information to confirm duplication. These duplicate responses were included in the qualitative analysis of open-ended questions but excluded from the descriptive statistics analysing project sustainability. The survey findings are integrated alongside the interview findings below.

Consistent with the evaluation questions outlined in our methods, the results are presented under three overarching themes: Impact, Implementation, and Sustainability. Our analysis generated 16 sub-themes, which are summarised in Table 2. Additional illustrative quotes for each sub-theme are provided in Appendix 4.

**Table 2.**
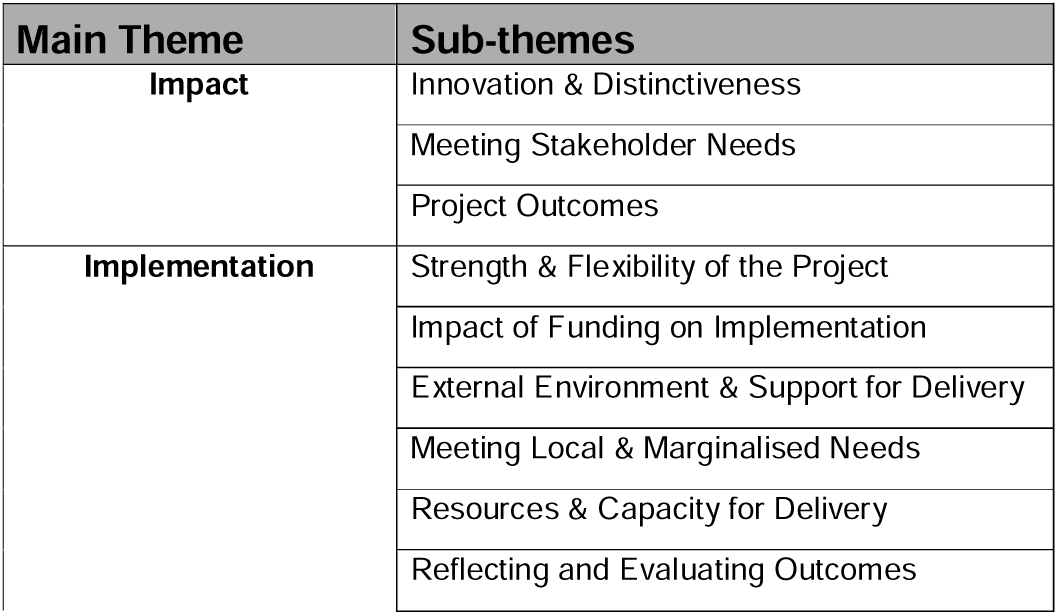
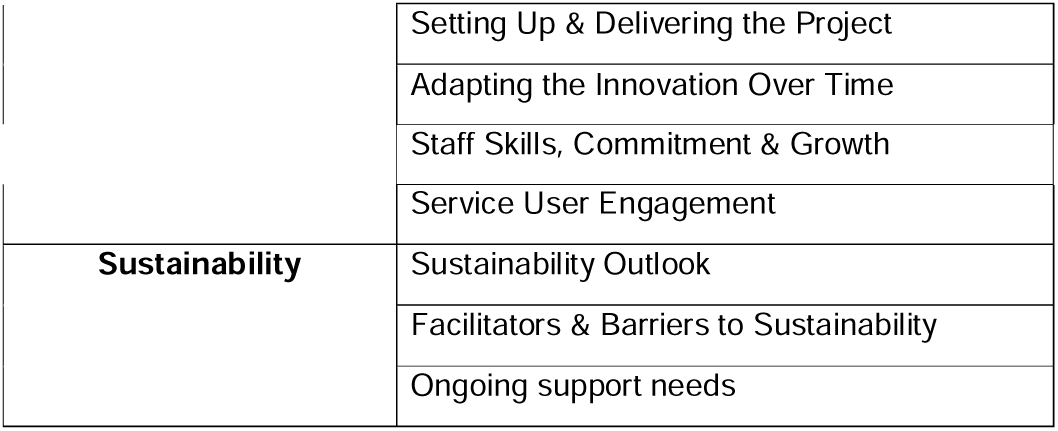
Sub-themes mapped to main themes.

## Impact

### 1. Innovation and Distinctiveness

The benefit of new funding was often in allowing projects to be set up that stood out as distinctive, either because of their novel features or status as a unique or rare service locally, regionally, or nationally. Often, they were reported to address important gaps in existing suicide prevention services. This included work with populations who are rarely targeted by mainstream services, such as burn survivors, children bereaved by suicide, or families of prisoners. At the same time, some emphasised the non-clinical, community-based or culturally adapted nature of their project as distinguishing them from regular NHS commissioning. As the participants noted:

*“The intervention that we deliver is not delivered anywhere else. It’s our own intervention that we’ve delivered and developed.”*

— *Participant 17b, Clinical lead*

*“There’s nothing similar really in the area, and there was a lot of feedback saying that people really wanted that help in those particular times.”*

— *Participant 7, Project manager*

### 2. Meeting Stakeholder Needs

Through this theme, we identified ways in which the projects meet the needs of both service users and other stakeholders, including service staff. The organisations demonstrated a strong commitment to meet the often-complex needs of their service users, perhaps through assessment, culturally appropriate intervention or trauma-informed models. For example, one interview participant explained how their programme was “absolutely” targeted at people at risk of suicide and included personalised goals ranging from “playing with their children” to simply leaving the house for the first time in years.

Some survey respondents indicated that they had consulted with relevant groups during planning (e.g., parents, young people), to tailor their approaches, whilst others mentioned operating flexible hours outside of 9am-5pm to ensure accessibility.

*“…we ran consultations with young people to see what activities they would like to see take place" - Survey 51, Project Lead*

The needs of staff and professionals were also taken into consideration. Many services adopted a whole-system approach, where they incorporated clinical supervision, debrief spaces, or professional training to equip staff to respond to suicide risk.

*“[The intervention] is a 24-hour suicide prevention helpline for anyone up to age 35 […] and also a debrief service for professionals and caregivers. We delivered four types of training depending on audience and need.”*

— *Participant 20, Contracts and finance manager*

### 3. Project Outcomes

Across interview participants, there was a strong belief in their project’s effectiveness, with many reporting significant improvements in well-being, coping, and emotional regulation among service users. Some services tracked measurable outcomes, including reduced anxiety and depression, improved quality of life, and higher-than-anticipated referral numbers.

*“Over 2024, we saw an increase of 42% in client self-esteem, and that was using the Rosenberg self-esteem scales. And then the GAD-7 scales showed a 58% reduction of anxiety levels, and we used the PHQ-9 scales, which showed the severity of depression decreasing by 57%. So together […] we saw an overall increase in client well-being of 52% over the course of 2024.”*

— *Participant 7, Project manager*

Most survey participants echoed this belief in their project’s effectiveness of tackling suicide rates and suicidality, mental health symptoms or wellbeing, and reaching large numbers of service-users. Only one explicitly identified their project as unsuccessful. Some additional insights were reported regarding improvement of social outcomes such as reduced social isolation and improved financial situations, as well as outcomes related to raising awareness of topics that allowed for earlier identification of people in need, or of their service which increased referrals and enabled more people to find support. Although these findings were reported, the specific measures employed were not clearly described in several survey responses.

*“For the local population, the benefits included reduced isolation, improved mental health and wellbeing scores, greater social confidence, and increased access to creative and supportive community activities. Members reported feeling safer, more connected, and more purposeful, with some developing employable skills and re-engaging in creative practice”*

— *Survey 52, Project manager*

### Implementation

We identified 10 sub-themes which mapped on to different CFIR domains (outlined below in Table 3). These are described in detail below, with illustrative quotes.

**Table 3.** Organisation of implementation sub-themes within CFIR domains.

| CFIR Domain | Sub-theme |
| --- | --- |
| Intervention Characteristics | Strength & Flexibility of the Project |
| Outer Setting | Impact of Funding on Implementation |
| Outer Setting | External Environment & Support for Delivery |
| Outer Setting | Meeting Local & Marginalised Needs |
| Inner Setting | Resources & Capacity for Delivery |
| Implementation Process | Reflecting and Evaluating Outcomes |
| Implementation Process | Setting Up & Delivering the Project |
| Implementation Process | Adapting the Innovation Over Time |
| Individuals | Staff Skills, Commitment & Growth |
| Individuals | Service User Engagement |

### 1. Strength & Flexibility of the Project

More than half of interview participants described their projects as robust and adaptable. Service managers highlighted that often their interventions were grounded in approaches they had previously delivered with success. In some cases, participants mentioned independent or academic evaluations that further strengthened their credibility among professionals and service users.

*“We weren’t necessarily doing anything brand new […] the University of Durham had done a report on the pilot project in the past.”*

— *Participant 2, Business director*

Others perceived that their longstanding delivery had earned them credibility and respect within their communities.

*“We have been in existence for 15 years […] we are highly respected within the region.”*

— *Participant 14, Founder and clinical director of the charity*

One survey response highlighted the strength of their online e-learning platform which had been subsequently incorporated into standard practice in their local NHS trust.

*“There was much interest from professionals working on the neurodevelopmental pathway in the quality of the course […] The elearning is now a standardised recommended course for parents on the [NHS Trust] neurodevelopmental pathway”*

— *Survey 2, Operational Lead*

Flexibility was another strength. These interventions were often co-designed with input from lived experience and tailored to the needs of the service users.

*“Our service is very flexible and tries to be quite comprehensive, […] we’re flexible to what the young person needs. […] we could meet them wherever they are*.

— *Participant 4, Project lead*

### 2. Impact of Funding on Implementation

Participants described the SPGF as essential to the implementation of their project. Several organisations noted that the funding and its flexibility allowed them to move beyond their existing services and focus on suicide prevention. Participants noted:

*“Without the funding, there’s no way that we would have had any kind of impact because we have to concentrate on the day-to-day, which is answering the phone.”*

— *Participant 15, Branch secretary and fundraising coordinator*

*“The flexible approach to reallocating funds as the project progressed, to meet emerging need, was super”*

— *Survey 3, Project Director*

The funding, for some, eliminated the administrative burden of having to apply for different funding throughout that period and therefore allowed them to focus on implementing suicide prevention.

*“Absolutely superb because it has removed the administrative burdens that we sometimes find and meant that we could really focus on frontline delivery.”*

— *Participant 2, Business director*

However, we identified several challenges in project delivery that were directly linked to the nature of the funding. Notably, participants across interviews and surveys highlighted the short duration (just over 1 year) of the grant as a significant constraint. This limited timeframe impacted their ability to develop sustainable interventions, recruit and retain suitable staff, and establish long-term, trusting relationships with target communities. As one participant noted:

*“ I think it was a relatively short time period to achieve what we did, and that was a challenge […] It was a very fast-paced approach that the staff had to take… [developing the intervention] can take two weeks or two years. You can’t force anybody or hurry anybody along.”*

— *Participant 6b, Project manager*

The same funding model was experienced differently, depending on the organisation’s size and capacity. Smaller charities perceived the administrative and reporting demands as disproportionately high for the scale of funding provided and found the bureaucracy burdensome:

*“There is an enormous amount of paperwork […] It takes up an enormous amount of administrative time and effort.”*

— *Participant 14, Clinical director and founder*

### 3. External Environment and Support for Delivery

This theme represents the wider picture as in the external environment surrounding the project and affecting the implementation. This could include local beliefs and culture around suicide prevention, policy or funding pressures, and external partnerships or networks that supported or shaped implementation. For example, the participants reported partnerships that were essential in getting the project started:

*“To getting the expansion started well, probably the most important was us making connections to the paediatric liaison nurses […] and the psychiatric teams that are based in hospitals […] so that they can refer young people that their teams have seen in hospital if appropriate, to our service. Because […] that’s where we got more referrals through […].”*

— *Participant 19, Project manager and company director*

Similarly to interview participants, survey respondents agreed that where these collaborations with local and third sector organisations worked, they notably helped with delivery, referrals and improving inclusivity. Novelly, whilst they identified positive experiences specifically in regard to NHS services, poorer experiences were mentioned relating to criminal justice services, such as prison staff being too stretched to provide logistical support, and schools, where lack of communication and buy-in were significant barriers.

*“we were welcomed by the NHS teams and the collaboration worked well”*

— *Survey 14, Senior client service manager*

*“Overcoming prison vetting regimes was an issue, as it took a while to get some workers into the custody environment”*

— *Survey 3, Director*

*“Our main challenges were around getting by in and engagement from schools and other organisations.”*

— *Survey 8, Overall lead*

Changes in the external environment, including political shifts and changes in government, created uncertainty around the project’s future. For some, these broader changes made the project feel less visible or less of a priority within the wider system.

*“[During active delivery] the government implemented what was called the early release scheme to tackle the huge overcrowding in prisons at that time. So, that meant there were a lot of changes within the prison. Lots of people get moved about and it creates a real atmosphere of uncertainty amongst the staff, […] because there was a lot of that, all the stuff going on, it was difficult to get the figures that we needed to evaluate effectively.”*

— *Participant 19, Project manager and company director*

Some of the criticism directed at the grant also speaks to a wider issue around how funding generally works for voluntary sector organisations. Fragmentation and duplication of funding were common concerns.

*“I do think the whole thing of charity funding needs a big, big rethink. I also think there are far too many very small charities. […] Now, logically and sensibly, they should all get together. And they should pool their resources. They’re all doing essentially the same thing.”*

— *Participant 14, Clinical director and founder*

*“I am also not sure about the merits of making so many small grants to so many organisations.”*

— *Survey 31, Director of data science, AI and research*

### 4. Meeting Local and Marginalised Needs

Service managers highlighted the project’s strong focus on local needs and their efforts to reach marginalised groups. Participants described working with marginalised groups over both survey and interview responses, including services offering free therapy or those delivered through prison settings were accessible to a wide range of individuals, meaning that marginalised communities engaged even when they were not the intended focus. This included work with refugees, asylum seekers, LGBTQ+ individuals, people experiencing poverty, those bereaved by suicide, and individuals facing multiple layers of social exclusion. Interviews found that many projects may not have targeted marginalised groups specifically, but nonetheless incorporated inclusivity throughout their offer to meet the needs of many local groups.

*“I think our objective was to go out there and to support everybody […] we’ve had people approach us from the transgender community. We’ve had young people come and talk to us. We’ve spoken to the homeless community. We’ve been out to food banks and spoke to people who are working but involved in the […] cost of living crisis.”*

— *Participant 15, Branch secretary and fundraising coordinator*

### 5. Resources and Capacity for Delivery

This theme captures the diversity of organisational structures and the various ways capacity was developed, stretched, or strategically managed to support suicide prevention. According to the participants, the capacity to deliver the project largely depended on staffing and infrastructure. Trends in the survey responses indicated that services which were already running and had an existing plan for outreach and expansion reported fewer challenges, whereas projects set up from scratch needed more time for set up and delivery.

*“The platform was already running before the project began, and we experience considerable demand, so there were no new challenges”*

— *Survey 31, Director*

Having clear roles and a chain of command was cited as essential to project delivery. The SPGF allowed for retaining existing staff and hiring new personnel to support activities.

*“The therapist would see the patients, and then the two supervisors who see the therapists. And then there’s a clinical lead […] So there’s like quite a clear chain of command for that.”*

— *Participant 10, Project manager*

*“I think it was [the funding] sufficient for sure. It was the right amount. I don’t think it was too much or too little. I think we estimated it pretty well.”*

— *Participant 6a, Head of trust and foundations*

Many organisations drew on the workforce of volunteers and external clinicians. Most described the funding as sufficient for delivery; however, one service manager mentioned venue availability as a key frustration, and another participant described safeguarding as a significant logistical challenge, stemming from a lack of staff.

“*Safeguarding was a massive thing, especially because we didn’t have a lot of staff. So, you would always have to make sure that there were at least two people in the building […].”*

— *Participant 7, Project manager*

### 6. Reflecting and Evaluating Outcomes

Participants described a range of approaches used to assess the effectiveness of their interventions and capture service user feedback. Standard methods included standardised clinical tools such as the PHQ-9 (diagnostic tool measuring severity of depression symptoms) [21], GAD-7 (measures severity of generalised anxiety disorder) [22], and CORE-10 (10-item measure of psychological distress across symptoms, functioning, and risk) [23] to track changes in psychological distress, well-being, and suicidal ideation, alongside qualitative and quantitative feedback forms. These were typically administered before, during, and after the intervention, providing insight into service user experience and perceived impact. However, in some cases, the participants did not clearly detail the specific nature of the feedback mechanisms used, or they were unable to capture service user feedback due to the anonymous nature of the service itself. Overall, participants valued feedback for accountability, reporting, service improvement and adaptation.

*“We use the […] Patient experience evaluation, and we use the best scale for suicidal ideation, PHQ-9 and GAD-7. We record outcomes at regular points throughout every survivor’s intervention with us, and they’re recorded on our patient database and then analysed each year.”*

— *Participant 1, Chief executive of the charity*

*“Parents were not using the elearning as we had anticipated and, instead, were using it as an encyclopedia type resource […] and not following through to the feedback section. This meant we had to pivot very quickly to a star rating at the end of each section”*

— *Survey 2, Operational lead*

### 7. Setting Up and Delivering the Project

We captured both the confusion in communication and the delayed clarity around funding. One participant reported *“a seven-month delay in us receiving the funding”.* This then caused a significant financial setback for the organisation.

*“By then, I had £90,000 left in my charity bank account, which was massively under where our operating funds should be.”*

— *Participant 5, Chief executive officer*

In response to delays in communication regarding their successful application, one participant explained:

*“What we needed to do was compress our already quite ambitious agenda, our ambitious objectives into a shorter space of time”*

— *Participant 6a, Head of trust and foundations*

Despite these early frustrations, participants consistently noted that once contact with the funding team was established, communication and support improved significantly. The efforts of named individuals were appreciated and frequently praised across interviews and survey responses:

*“But then once we had the grant manager […] in place, she was way more communicative.”*

— *Participant 10, Project manager*

*“Our lead contact in DHSC was always so helpful and pragmatic. It really made a difference to the success of the project and our ability to ’get on and deliver”*

— *Survey 3, Project Director*

This turning point was widely described as a key facilitator in getting delivery on track.

### 8. Adapting the Innovation Over Time

The staff often made changes to the intervention, due to a need for further training, changing priorities, or emerging needs from feedback from service users. This theme clearly reflects an effort and a desire to meet service user needs.

*“We started getting quite a few requests from parents and carers for training and support for them. And we don’t offer like, you know, intervention with adults. But what we started […] to do regular trainings with parents and carers and also […] other people who support young people in mental health crisis. So, it could be like a teacher or you know, community nurse or something.”*

— *Participant 4, Project lead*

Some survey respondents shared that they had adapted their services in response to challenges by diverting funds away from areas that were proving difficult to set up to focus on delivery, by reallocating already vetted staff to combat vetting challenges, or by combining multiple aspects of their project into one resource to be more cost effective.

*“We struggled with recruitment of our specialist counsellor, so our expected start time for this support was delayed by a few months […] and we diverted funds from this part of the project into the other therapeutic activities so that we could continue benefiting survivors.”*

— *Survey 13, Administrator, monitoring and collating data*

In addition to the adaptations already made, participants also shared suggestions for improvements they would make if they were to redesign the project. Their responses highlighted a strong desire for more collaboration and greater visibility for the work through improved media engagement.

*“Because I think the social media and probably promotion of the funding were maybe a bit weak. Not from our end, but from the DHSC […] I think it should be something we should be shouting about, and the fact that the government invested such a significant amount of money shouldn’t be lost, and I think it was. Now we obviously shouted about that ourselves, but again, coming back to the shared experience, the peer support, the collaboration and I think that was missed.”*

— *Participant 20, Contracts and finance manager*

### 9. Staff Skills, Commitment and Growth

Around two thirds of interview participants reflected on the knowledge, skills, and motivation of staff. They described their teams as empathetic and committed to creating safe spaces for vulnerable individuals. Staff training was at the forefront for all organisations, especially in suicide prevention and safeguarding.

*“We have specific training for staff on suicide prevention. And again, that is a regular thing that we will be pushing due to staying at the forefront of learning and understanding.”*

— *Participant 3, Head of counselling and well-being*

The recruitment was taken seriously, with onboarding often taking a long time, making sure the staff was fully prepared for delivering the intervention.

*“It takes us three months to train one person to the level they need to be at to be able to identify, negotiate and then remove that person, and then even then, they’re three months in probation.”*

— *Participant 5, Chief executive officer*

Some survey respondents had recruited lived experience staff and volunteers, staff from diverse backgrounds, and qualified staff with specialist experience to ensure they provided high quality, culturally competent support.

*“[We] recruited 5 peer support volunteers with lived experience of bereavement related to postpartum psychosis. This group supported [us] to develop targeted resources for bereaved families and supported the delivery of specialised peer support for affected families. [Our] staff attended a range of training courses related to grief and suicide prevention to strengthen their knowledge and skills in these areas.”*

— *Survey 47, UK Programme Manager*

Overall, these responses reflect that organisations were cautious and appropriately skilled in developing and delivering their interventions.

### 10. Service User Engagement

Less than half of our interview participants reflected directly on service user engagement. These reports expressed mixed experiences with some noted strong uptake, while others struggled to recruit service users. This feedback was similarly mixed in feedback from the survey respondents:

*“The group sustained a solid core membership of eight or nine people who came regularly, nearly every week. […] it was immediately taken up; the group was immediately subscribed to and people immediately started using it in the way that they did.”*

— *Participant 8,* Programme manager/peer group facilitator

*“I think that’s been the hardest thing when you’re running an event and inviting people to come to give them a reason and to get the numbers. And that was tough.”*

— *Participant 15, Branch secretary and fundraising coordinator*

*“It was hard to get parents / carers to participate in face to face or online training in spite of high levels of promotion”*

— *Survey 36, Lead professional*

*“Activities for Neurodiverse/LGBTQ+ were extremely popular and always to capacity.”*

— *Survey 51, Project lead*

One participant spoke of reluctance and apprehension early on, which turned into deep involvement as time went by:

*“[They] turn up a bit scared and apprehensive, thinking, what’s this? Who are these people? What am I signing up for? They then go on to pretty much run groups and share the skills they have.”*

— *Participant 7, Project manager*

## Sustainability

### 1. Sustainability Outlook

Across the interviews, participants expressed mixed outlooks regarding the future of their funded interventions. Several reported that their projects would cease entirely once the SPGF ended, primarily due to a lack of continued funding. Others indicated that the work would partially continue, albeit at a reduced scale or with fewer of the original components.

*“It will [continue] if we can, but like every charity, we can’t do it unless we’ve got the funding, and it’s a huge amount of funding for us to replace.”*

— *Participant 1, Chief executive of the charity*

In a closed survey question, respondents were asked “Is the project going to continue now that the funding period is coming to an end?” Out of 29 eligible responses, 41.4% said yes, 20.7% said no and 37.9% said that they were unsure whether the project could continue. When given the opportunity to expand on this, they had similar concerns over facing closure, scaling down projects, and inability to retain trained staff.

*“The lasting affects of training will be embedded into our practice, but we are unable to afford new staff being trained in this level.”*

— *Survey 6, Project Manager*

Only a minority of participants stated that their intervention was fully sustainable beyond the funding period; these tended to be digital resources (e.g., websites) that require minimal ongoing costs, services that developed partnerships during the funding period, or interventions that were already embedded within existing, well-established services, enabling them to continue as part of routine provision.

*“Our project is not really time bound. […] the groups that we’ve developed will continue on, probably for years after the funding.”*

— *Participant 6a, Head of trusts and foundations*

*“Partnerships established as part of the pilot have continued after project completion, which will ensure that we can continue to deliver the workshops within their establishments reaching more students and improving their wellbeing and increasing their ability to achieve academically.”*

— *Survey 33, Project Supervisor*

Survey responses showed that the motivation to continue projects at a reduced scale and prioritise continuation despite the challenges was high.

### 2. Facilitators & Barriers to Sustainability

We identified barriers and facilitators to sustainability across interviews and survey responses. These are summarised below and in Table 4.

**Table 4.** Summary of the main barriers and facilitators to sustainability identified across interviews.

| Theme | Type | Quote (Paraphrased) |
| --- | --- | --- |
| Short-term funding | Barrier | Only 18 months of funding; staff left before project ended. |
| Lack of continuity funding | Barrier | No appetite from the funder for continuity funding despite demonstrated impact. |
| Poor communication with funder | Barrier | Communication with the funder was virtually non-existent after initial contact. |
| Reliance on fundraising | Barrier | Heavy reliance on public fundraising which is declining year by year. |
| Staff recruitment challenges | Barrier | Staff hesitant to commit to short-term roles without long-term prospects. |
| Political uncertainty | Barrier | Changes in NHS England and political context reduced likelihood of future funding. |
| Fragmented funding landscape | Barrier | Many small charities doing similar work due to fragmented, competitive funding. |
| Sustained external funding | Facilitator | Secured five-year lottery grant integrated into core programme. |
| Strategic financial planning | Facilitator | Developed centres with financial buffers to ensure medium-term operation. |
| Recognition by funders | Facilitator | DHSC funding improved credibility and helped attract new funders. |
| Ripple effect of stigma reduction | Facilitator | Challenging stigma has created a lasting cultural impact in prison settings. |
| Funding from other sources | Facilitator | Follow-on funding secured to extend project delivery beyond initial grant. |

**a)** Barriers

One key barrier was the short-term nature of funding, which several participants felt undermined the long-term impact of their work.

*“It was disappointing to learn that there wasn’t any continuity funding… it feels like we’ve demonstrated the impact we have.”*

— *Participant 1, Chief executive of the charity*

This barrier, in turn, relates to additional issues regarding staff retention, the inability to plan ahead, and insecurity about the project’s future.

*“I guess a lower level, like with it being only 18 months of funding, staff are aware of that when they take on the role. […] not having something long-term for staff when you’re recruiting for staff to be able to think, Oh, this might be a job I can stay at for a bit longer.’ That is also, I would say, difficult for an organisation.”*

— *Participant 16, Fundraising manager*

**b)** Facilitators

We identified several facilitators for sustainability. The most prevalent one was securing follow-on or external funding, like gaining lottery or local grants, or using DHSC funding to leverage new funding. Participants describe the SPGF as a “stamp of approval” from DHSC that helped establish the credibility of the organisation and positioned them as trustworthy in the eyes of future funders.

*“[…] getting the funding from the Department of Health and Social Care was great at leveraging funding since and helping us to continue to fund the service as we’ve talked about this stamp of approval that we’ve got. So, it’s really helped us engage new funders […].”*

— *Participant 17a, Fundraising manager*

### 3. Ongoing Support Needs

A near-universal theme among participants was the critical need for continued funding to ensure long-term sustainability. While some highlighted other support needs such as staffing, training, or partnership development, the availability of dedicated financial resources was consistently identified as the primary enabler for maintaining or scaling their project beyond the grant period.

*“I think we’re doing everything that we possibly can at the moment […] there’s literally not much else that we can do without funding […].”*

— *Participant 15, Branch secretary and fundraising coordinator*

In some cases, participants had clear ideas about the amount of funding needed and its appropriate form.

*“It’d be a grant, and I think it would be ideally in the region of about 500,000 pounds or more, just to be able to offer a long or even longer period of therapy. Ideally, it could be an element of unrestricted funding to cover core costs.”*

— *Participant 10, Project manager*

Several projects highlighted the value of the SPGF and its processes, particularly its organisational and communication support, with strong support given for its continuation. However, a few highlighted need for increased flexibility around funding model and recognition of financial and reporting burdens on small organisations in future.

*“Communication from the DHSC has been very responsive and supportive, and payments received efficiently.”*

— *Survey 13, Project Administrator*

*“Understanding the cost implications of reporting in annual accounts/auditors is important when working with smaller charities.”*

*—Survey 16, CEO*

Additionally, participants expressed a need for the DHSC to be more involved with the funded projects, such as directing them towards future funding and even witnessing the intervention themselves.

*“It’s always difficult to describe an art intervention. So, it would be great if representatives from the Department of Social Care could actually come and sit in on a session and actually see and witness it.”*

— *Participant 19, Project manager and company director*

## Discussion

Our study explored the perceived impact, implementation, and sustainability of services funded through the SPGF. Our findings highlight some of the strengths and limitations of the funding.

An injection of time-limited funding allowed VCSE organisations to set up projects that participants widely regarded as distinctive and innovative, often filling critical service gaps and reaching groups underserved by mainstream provision (e.g., bereaved children, burn survivors, families of prisoners, LGBTQ+ communities). Many services reported improvements in well-being and coping, with some reporting reductions in distress, anxiety, and depression on routine outcome measurements. However, these findings should be interpreted with caution: establishing the impact of the funded projects on local suicide rates was beyond the aims of the SPGF and inherently hard to demonstrate in the timeframe and scale of the funded projects.

The funding was essential for delivery, enabling organisations to expand or establish suicide prevention initiatives. Key enablers of implementation included flexible, co-designed approaches, skilled and motivated staff, and productive partnerships with local systems. However, organisations also reported barriers to implementation, particularly short timescales for delivery, high administrative burdens, and difficulties engaging some service users.

Sustainability of interventions was the most significant concern for managers. While some interventions (primarily digital or embedded services) are expected to continue, most projects faced uncertainty or closure once SPGF funding ended. The short-term nature of the grant limited capacity for long-term planning and staff retention. Nonetheless, SPGF provided a “stamp of approval” that enhanced credibility and helped some organisations secure follow-on funding.

Overall, while service managers saw the SPGF as vital for delivery, its structure and limitations posed significant barriers to full and sustainable implementation. Figure 1 summaries our main findings and how implementation barriers and facilitators impact intervention sustainability.

**Figure 1.**
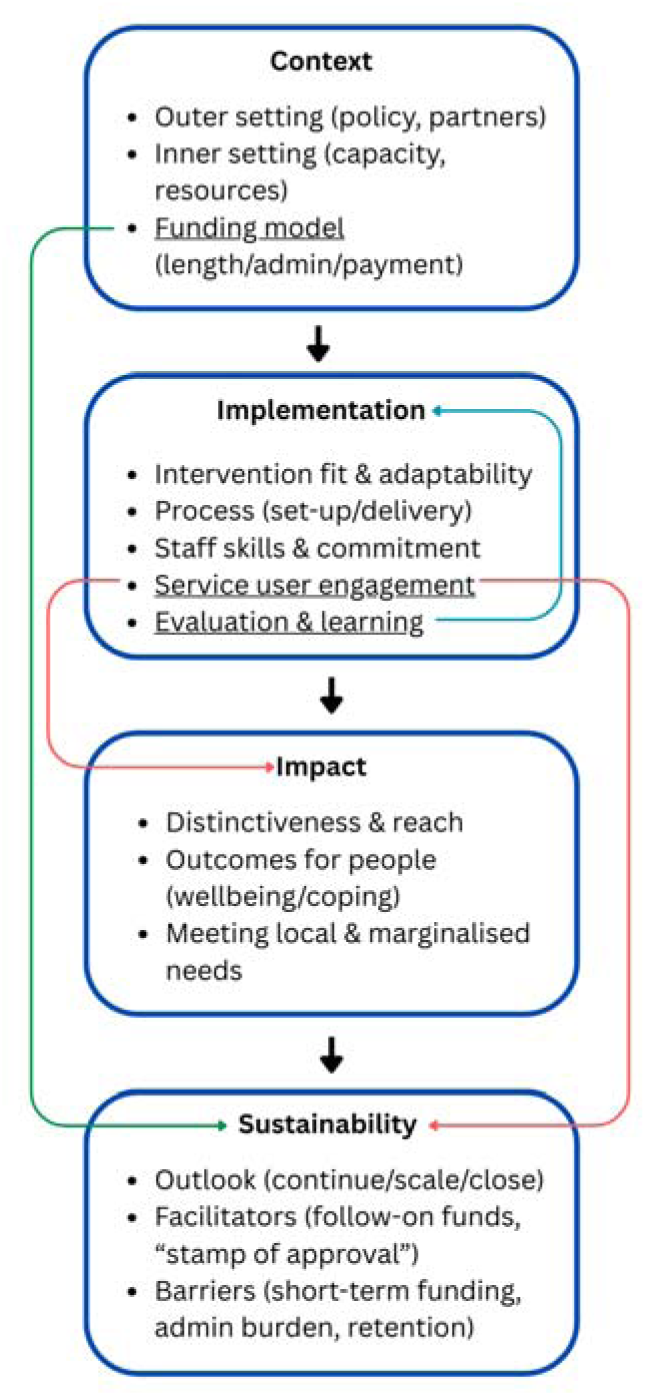
Diagram illustrating the relationships between findings.

*Outer/inner setting and funding model shape implementation processes, which drive impact; evaluation creates feedback loops that inform adaptation; engagement and funding structures directly affect sustainability and equity*.

### Limitations of the study

Several limitations should be considered. Recall bias may have influenced participants’ views due to the time between project setup and interviews. Additionally, only interviewing service managers involved in delivery might have introduced positive bias, as their strong involvement could lead them to emphasise strengths, possibly excluding less satisfied perspectives on outcomes. The SPGF was awarded to 78 projects, and we interviewed 20 of them, which was our initial goal. We expanded the scope to attempt to capture important perspectives that we may have missed by including the additional short survey. The survey responses, whilst providing additional nuance to the findings from the interviews, did lack the same depth as interviews. The interviews captured a mixture of positive and more critical reflections, suggesting that we captured a range of in-depth views. Interviews were conducted soon after funding ended, limiting insights into long-term impacts or sustainability beyond June 2025. Future follow-up, at later points, could reveal if and how the SPGF enabled projects to endure beyond the grant.

Importantly, the study was limited to exploring service managers’ perceptions of impact rather than directly measuring the effectiveness in preventing suicide. Whilst some managers reported improvements in mental health, wellbeing and other social outcomes, we are unable to draw firm conclusions on the effectiveness of the included interventions on reducing suicide rates.

### Challenges of the Grant Fund

The responses show the grant fund was impactful in supporting vulnerable groups at risk of suicide, helping every interviewed service meet their core aims. However, there was criticism about the short-term funding, seen as a major barrier to sustainability. Participants stressed that without ongoing investment, momentum and outcomes could be lost. Uncertainty about future funding caused staff to feel unsure, disrupted long-term planning, and sometimes led to early scaling back of successful initiatives.

Smaller charities found the retrospective payment structure burdensome, as they lacked the reserves to cover upfront costs. While larger, well-established organisations generally coped better, this raises concerns about equity: the very charities most embedded in marginalised communities may be the least able to access or fully benefit from such funding.

Additionally, there was a shared view that the visibility and promotion of the fund could have been stronger. Some participants suggested that the DHSC, being a respected and authoritative body, should play a more active role in signposting organisations toward longer-term funding opportunities and advocating for the continuation of successful projects.

### Comparison with previous research

Our findings align with previous research on the voluntary sector in mental health and suicide prevention. Baxter and Fancourt (2020) [24] observed that voluntary organisations are often established based on lived experiences and gaps in services, but they face ongoing funding insecurity.

Similarly, Newbigging et al. (2020) [25] highlighted the relational and peer-based qualities, such as kindness and compassion, of VSCE organisations as central to their effectiveness. These qualities clearly echoed in our participants’ accounts of service users later becoming volunteers or staff.

As noted earlier, smaller charities often struggled with the retrospective payment structure and the short-term nature of the grant. Additionally, some projects also reported difficulties in sustaining service user engagement. Taken together, these barriers suggest a risk that the SPGF inadvertently excludes the very “under the radar” charities that are best placed to reach marginalised communities. This concern was also raised by Newbigging et al. (2020), and it is something that should be carefully considered by funding bodies when designing future funding opportunities of this kind.

Other national funding programmes, such as the Better Care Fund evaluation [26], similarly highlighted that implementation is shaped as much by organisational and contextual barriers as by the funding itself. Although the Better Care Fund focused on statutory health and social care services, and the SPGF targeted voluntary and community organisations, both point to the importance of strong leadership, shared vision, and sustainable structures to translate funding into long-term change. Additionally, a novel systematic review [27] identified key implementation determinants using the CFIR framework, such as leadership engagement, resources, compatibility, and relative advantage, reflecting our sub-themes like capacity, adaptability, and external support.

### Implications for policy and practice

Our findings highlight key implications for policymakers. While short-term grants foster innovation, longer funding cycles are recommended for lasting impact in suicide prevention. Continuity investment for successful projects is likely to reduce turnover of trained staff, maintain relationships between local services and help provide consistent service provision, leading to improved sustainability. In the SPGF, funding ended after a year regardless of success, leading to closures or scaling down of potentially effective projects. Up-front payments, instead of retrospective reimbursements, would enable smaller organisations to participate more equitably. Better coordination among charities could reduce duplication and fragmentation, expanding support networks. Additionally, promoting funded projects can share best practices and highlight the voluntary sector’s role in national suicide prevention. Lastly, stronger partnerships between government, statutory services, and VCSE organisations could embed successful interventions into wider care systems, reducing the risk of losing effective projects after funding ends.

## Conclusion

This study provides the first independent evaluation of the SPGF, highlighting both the opportunities and challenges of funding VCSE organisations in suicide prevention. Our findings suggest that short-term funding can increase the capacity of voluntary sector organisations to deliver suicide prevention activities, particularly in areas with high levels of marginalisation and unmet need.

The findings contribute to ongoing debates about sustainability in the voluntary sector and inform policy development around future funding models, both within and beyond the context of suicide prevention.

### Lived Experience Commentary

Karen Machin and Kathleen Fraser

Whilst timelines precluded full collaboration in this evaluation, we joined planning meetings, and were involved in analysing transcripts and identifying themes. Our input was informed by our personal experiences not only in relation to the topic of suicide, but also of working, volunteering, and being supported by small organisations and projects within the voluntary sector.

We know that these organisations “meet people where they are”: they are flexible and have values including co-production which are important to people who seek their support. We were impressed to see acknowledgement of this in the findings of this study.

We also welcomed corroborations of views long expressed by carers such as “prison staff being too stretched”. Prison is not the best place for people with mental health needs. The pace required for permissions and access in prisons requires longer term consideration, not short-term projects.

This paper offers some suggestions on how funders might develop their approaches and encourages a move away from short-term funding. For example, provision of IT tools/templates could ease the disproportionate reporting that burdens smaller responsive organisations.

We question if future funding should be geared to innovation or to filling gaps in existing services. Short-term funding of projects can provide an exciting opportunity to try out new approaches, but it clearly leads to issues in terms of recruiting, training and retaining staff. As people looking for support, it is disheartening to see valued projects come to an end and trusted, reliable people move on. Sustainability of projects is the minimum we are looking for: ideally we would like innovative projects to develop further.

We disagree with the quoted comment that too many small organisations are doing the same thing. We know they are different. We value the innovation and relational working of smaller organisations who respond swiftly to local community needs. The additional funds, and this paper, offer useful feedback about supporting these smaller organisations.

## Declarations

### Ethics approval and consent to participate

Ethical approval was not required as this study was classified as a service evaluation. All interview participants provided informed written consent prior to taking part in an interview.

### Consent for publication

Not applicable.

### Availability of data and materials

The datasets generated and/or analysed during the current study are not publicly available due to ethical considerations but are available from the corresponding author on reasonable request.

### Competing interests

Authors JT, KA, RD, AG, PN, AS, SJ, RA, BLE are co-Investigators or employed research staff in the NIHR Policy Research Unit in Mental Health, which undertakes commissioned research for the English Department of Health and Social Care, who also funded the Suicide Prevention Grant Fund programme.

## Funding

This study is funded by the National Institute for Health and Care Research (NIHR) Policy Research Programme (grant no. PR-PRU-0916-22003). The views expressed are those of the author(s) and not necessarily those of the NIHR or the Department of Health and Social Care.

## Authors’ contributions

DV conducted all interviews and led data analysis, with supervision from RA and BLE. Survey data were collected and analysed by JT and KA. All authors contributed to the coding framework and interpretation of findings. The initial manuscript was drafted by DV, with input from JT, KA and RA. AG and PN oversaw the other lived experience researchers involved in the project. All authors read and approved the final manuscript.

## Data Availability

Anonymised data produced in the present study are available upon reasonable request to the authors

## Notes

### Author Declarations

Our protocol was reviewed by Noclor NHS Research Office, the R&D department of North London Foundation Trust on 2nd April 2024, in line with HRA guidance to seek R&D advice not REC advice for projects which will not be managed as research Noclor confirmed that the study meets criteria to be classified as a service evaluation.

### Summary of Updates

We have added a Lived Experience Commentary at the end of the paper. This is a brief reflective discussion of key issues relating to the study and its findings, from the perspectives of two authors with mental health lived experience. We include a Lived Experience Commentary with all papers connected to the NIHR Policy Research Unit in Mental Health, to provide an unmediated lived experience voice. The text of the paper is otherwise unchanged.

