## Supplementary material for "Evaluating the Impact, Implementation and Sustainability of the Suicide Prevention Grant Fund – A Qualitative Study"

### Appendix 1

**SPGF services for qualitative interviews: sampling grid**

| Target Population | Provider type | Bid type | Area – suicide rate | Area - region | Service type | Development type | Funding amount |
|---|---|---|---|---|---|---|---|
| a)Whole population<br>b) SPGF priority group<br>c) Other group | a) Mental health service<br>b) Other service | Consortium bid Y/N | a) High suicide area*<br>b) Lower suicide area*<br>c) Middle | Which English region | a) Clinical or educational<br>b) Other | a) New service<br>b) Addition to existing service | a) Small <10k<br>b) Medium 10-100k<br>c) Large >100k |
| b) burn<br>survivors | a) and b) | N | N/A | National | b) | b) | c) |
| b) people in contact with the criminal justice system | a) | N | a) | North East | a) | a) new service | c) |
| a) support for people with life-threatening conditions | a) cancer counselling | N | a) | South West | a) counselling service | b) | b) |
| b) CYP | a) | N | c) | South East | c) | b) | c) |
| a) | b) search and rescue | N | c) | South East | c) | b) | c) |
| b) men | community group | N | Various | North of England and East Midlands | a) community groups focusing on reducing social isolation | b) setting up new community spaces for men | c) |

| | | | | | | | |
|---|---|---|---|---|---|---|---|
| b) and c) | b) | N | C) | Hertfordshire | educational | a) piloting out-of-hours service | c) |
| b) includes people with previous suicide attempts | a) | N | a) | North West | b) peer support groups | b) | a) |
| c) people with autism | b) | N | a) | North East | a) Producing toolkit and web platform | b) new platform and toolkit | c) |
| a) | a) | N | b) | London | a) (screening SP videos) and b) clinical treatment | a) new Suicide Prevention Service | c) |
| b) CYP | b) | N | a) | South West England | a) | b) | a) |
| b) CYP | a) | N | a) | South West | a) | a) and b) | b) |
| b) men | a) | Y | b) | London | a) | a) and b) | b) |
| a) | a) | N | a) | North West | b) | b) | b) |
| a) | a) | N | a) | Yorkshire and Humber | a) and b) | b) | a) |
| b) CYP | a) | N | b) | South London | b) | b) 'extension and reconfiguration of existing work' | c) |
| b) men | a) | N | a) and c) | South East, North East and North West | a) | b) expanded locations of their service | c) |
| c) women with previous MH contact | a) | N | a) | Yorkshire and Humber | b) | b) continuing long-term trauma therapy service | b) |

| | | | | | | | |
|---|---|---|---|---|---|---|---|
| b) men in contact with the criminal justice system | b) | N | N/A | Based in North West but the service is national | c) Interactive theatre performances | b) expansion of a previously piloted intervention | b) |
| c) under 35s | a) | N | N/A | National | a) and b) | a) creation of community hubs, supported by both SPGF and organisational funds | c) |

*High suicide rate areas = North East, North West, Yorkshire, South West; Low suicide rate = London Suicides in England and Wales - Office for National Statistics

### Appendix 2

### SPGF Qualitative Evaluation Project

#### Topic Guide

**Participant information**

1. To begin, I would like to ask you a few background questions about your role and experience, if that is alright with you.

- Your role on the SPGF-funded project
- How long have you worked in your current job?
- How long have you worked in roles related to mental health or suicide prevention?

**SECTION 1: Perceived Impact**

2. Please can you briefly describe the project which was funded through the Suicide Prevention Grant Fund?

- What were the aims of this project?
- Was this a brand-new initiative, or an expansion of an existing project or service?

If expansion:

- What was the rationale for expanding the initial project? (eg, to meet the extra demand or to market the service to more people)

3. In what ways was your project innovative or different from existing approaches - either within your organisation or more widely?

- Was there anything new or distinctive about the type of service you provided?
- Was there anything new or distinctive about how you delivered your service?
- Who staffs the project, and how do they relate to the wider team or organisation?

- Was your project based on an existing model of support? What outcomes were you hoping for when you applied for funding?

4. How did you hear back from your clients or service users about the project?

- Did you collect any feedback on people's experiences with the project? Are there any changes you've made or plan to make because of what you heard? If yes, please elaborate.

**SECTION 2: Implementation**

5. How did you find the process of applying for and setting up your SPGF-funded project?

- Were there any challenges or aspects that you think could have been improved in the grant process?

6. How has the SPGF-funded project impacted your service or local suicide prevention efforts?

- Can you give examples of any changes or impacts on outcomes that you and your team have noted? Positive or negative?
- Did the community or service users benefit from the funding? If yes, how? If not, why not?
- Do you feel that the funding helped you meet the specific needs of your local population?
- Were you able to support any particular marginalised groups?

7. What helped with delivering your SPGF-funded project?

- Were there any partnerships or collaborations that were important to getting the project started?
- Did you work with any other local services or organisations — including NHS services — to help deliver the project? If yes, what role did those partners play in helping you get started?

8. What if anything made it difficult to deliver your SPGF project?

- If any challenges mentioned: how did you overcome these?
- Did you have the resources (training, staff, time, space) you needed to implement the project effectively?
- Did you need to recruit any additional staff to deliver the project?
- If yes, what was your experience with recruitment or retention?
- Did the available funding feel sufficient for what you were aiming to do?

9. Do you think the project achieved what you originally set out to do?

- Were there any outcomes you didn't expect initially?
- Did anything change over time — such as unexpected barriers or shifts in priorities?
- If yes, how did you overcome these challenges?
- Was there anything that took more time or effort than you expected?
- Is there something that worked particularly well that you'd repeat?

**SECTION 3: Sustainability**

10. Is the project going to continue now that the funding period is coming to an end?

- If yes, what's making it possible for the project to continue?
- If not, what are the barriers to continuing the project?

11. What would help the sustainability of these projects in the long term?

- Is there potential to make it part of standard practice or the usual activities in your service?
- What kind of funding would be needed to keep the project going?

**SECTION 4: Ending the interview**

12. Is there anything you would do differently if you were going to set up this project again?

13. Is there anything else which we haven't asked you about yet that you would like to mention?

**Thank you so much for your time and insights today. They've been really valuable.**

### Appendix 3

#### SPGF Qualitative Evaluation Project

##### Survey Items

Respondent Profile Items:

1. Name of SPGF-funded project
2. Your name
3. Your role on the project

Survey Items:

1. Please can you briefly describe the project which was funded through the Suicide Prevention Grant Fund and its aims?
2. What were the impacts of the project? e.g. any changes to your service delivery, any benefits for your local population?
3. What helped and what were the challenges with setting up and running the SPGF project?
4. Is the project going to continue now that the funding period is coming to an end?
    a. If yes, what is making it possible for the project to continue?
    b. If no, what are the barriers to continuing the project?
    c. If the project might continue in some form or if you're currently unsure whether it will continue, can you give us some more details?
5. Do you have any suggestions for improvements to how funding schemes like the SPGF are set up and organised in future?
6. Is there anything else you'd like to mention?

### Appendix 4

#### Sub-themes mapped to main themes with illustrative quotes.

| Main Theme | Sub-themes | Interview Quote | Survey Quote |
|---|---|---|---|
| **Impact** | Innovation & Distinctiveness | *"I think the peer-support element is innovative in that it emphasises people's assets and strengths in order to get them through difficult times, whereas mainstream support is often based on a deficit model [...]. (P12)"* | ***"There was much interest from professionals working on the neurodevelopmental pathway in the quality of the course, the new approach to reaching parents and the volume of parents reached in a short period of time [...] is now a standardised recommended course for parents on the North Lancashire neurodevelopmental pathway" (S2)*** |
| | Meeting Stakeholder Needs | *"We offered training to people and to other organisations, so they knew how to communicate and look for signs of mental ill health in people with autism." (P13)* | *"We respond to increased demand for family support, providing support via a helpline and in courts, as well as support for the children of people in prison." (S3)* |
| | Project Outcomes | *"We really saw improvements in survivors and how they were coping. [...] we didn't lose any survivors to suicide, which feels like a really crude measure, but equally a very important one." (P1)* | "We were able to increase our team of specialist staff, reduce our waiting lists and work with more victims of abuse reducing their distress and risk of suicidal action." (S25) |
| **Implementation** | Strength & Flexibility of the Project | *1."We're here to support people's health and well-being, but we're a creative, fun space. We're a place where you don't have to talk about it if you don't want to, but we're here for it." (P6); 2. "[The intervention] is led by psychoanalysts. You have very in-depth knowledge of the idea of approaching suicidality." (P10)* | *"a cost effective, digital support service for one of the most at risk groups" (S4)* |
| | Impact of Funding on Implementation | *"We were asked for reports that just don't exist in the charity sector [...] three months after the financial year end. Charities don't have to publish their accounts for nine months. So again, that wasn't really a very realistic expectation." (P1)* | *"I feel our biggest challenge was time, 12 months is a very short time period in which to set up and run a project and engage professionals working in those fields to recognise it's value. However, it was a great opportunity to pilot something that we hadn't tried before and we wouldn't of had the monies to do so, despite us having the capability. We were just getting started on the elearning success essentially, when the monies were coming to an end." (S2)* |
| | External Environment & Support for Delivery | *1. "I guess at a time when there was lots of uncertainty in the government. Really. So maybe it wasn't something they wanted to prioritise [...]" (P1); 2. "We had a good relationship with local social prescribers, so that helped us get a lot of our initial clients. We worked closely with [...], a substance misuse provider in the area. So, we kind of partnered with them." (P7)* | *1* "The main challenges were around referral numbers with our projects [....] Good relationships with A&E nurses as key referrers was a key thing that helped." (S1)<br>**2** *"Challenges were getting local services to support and utilise this service." (S7)* |
| | Meeting Local & Marginalised Needs | *"We try to consistently offer our service in marginalised areas when we can [...] alongside that, we try to work as a team to be as well-trained in the EDI as possible." (P3)* | *"The funding [..] had a significant impact, enabling us to expand our capacity, clear a long waiting list, and **reach many new groups — including LGBTQ+ communities who had not previously engaged with the project** " (S52)* |
| | Resources & Capacity for Delivery | *"We have a fantastically high retention of staff. We recruit twice a year. We have huge amounts of support. We have a crisis counsellor on the team. We also have a team of supervisors, and we also have* | *"with a short 12 month project it is difficult to provide the ongoing work needed for those with mental health whilst also working towards the KPIs"* (S5) |

| | | | |
|---|---|---|---|
| | | *church support for every team member." (P5)* | |
| | Reflecting and Evaluating Outcomes | *1. "The service users complete screenings at the beginning, middle and end, they have screenings which are based on three different standardised questionnaires [...] the CORE-10, [...] then a service user questionnaire to understand what other services they're using and if they're using that by the end." (P10); 2. "It's completely inappropriate for us to be asking how they feel about our service. We're only with them from that window that they are in suicidal ideation. Once they've gone, we have no connection." (P5)* | 1 "Feedback from survivors and staff have been very positive and the impact significant. " (S13)<br><br>2 "Participating prisons have not released support service engagement figures post Man Up engagement, making it difficult to make solid determinations on the impact in this area." (S37) |
| | Setting Up & Delivering the Project | *"We found the process actually really straightforward, [...] Where we then had an issue was that there were big delays in awarding the project. So, there were delays, and it went on, and so we went past when we were supposed to have started." (P2)* | *1 "Setting up a new project, or in our case growing and developing an additional one is front loaded and time consuming e.g. recruitment and training so to have to end the project after 12 months feels really unsatisfactory on many levels." (S28)*<br><br>*2 "**we didn't face any challenges setting up or running.** we were welcomed by the NHS teams and the collaboration worked well" (S14)* |
| | Adapting the Innovation Over Time | *"If I'd had more time in the beginning [...] to do all the marketing, make all the contacts and maybe have [...] an opening event [...] it was slow starting off. [...] If you have that time in the beginning, you can do all your training before you open as well." (P7)* | *"We changed the 2 resources into a combined single pocket sized concertina booklet following a) increased printing costs b) feedback from prisoners." (S16)* |
| | Staff Skills, Commitment & Growth | *1. "Our training is very comprehensive. And it's kind of it feels like we have many fail safes to make sure that if there are safeguarding situations then they won't go unnoticed — and there often are." (P4); 2. "And so it was about bringing people in who either have lived experience of suicide directly or indirectly, and others who just have an interest in suicide and helping and mental health to give them ownership of some of what we do." (P20)* | *"The DHSC funding also allowed APP to support staff from diverse backgrounds to ensure culturally competent support is available to women and families from different ethnic groups. " (S47)* |
| | Service User Engagement | *"We want the people to come in every week. So, if you have something that stops that for a week or two, it's hard to get that momentum, particularly when it's been very difficult to get these people here in the first place." (P13)* | *"Our other challenge was around engagement. We had a high volume of non attendance on our face to face and virtual workshops. [..] This meant that a lot of our reliance on engagement relied on the success of the elearning which we felt may be a more accessible way for parents to engage in the training. " (S2)* |
| **Sustainability** | Sustainability Outlook | *"The project will automatically continue, and we've been around for 20 years, so this funding has guaranteed that we're financially stable till the end of 2025, and our funding strategy will guarantee us to move forward." (P5)* | *"**It's been great for us to finally be able to pilot this project, and now we have done this, we are in a great position to apply for longer term funding** (great positive feedback, data regarding activities and numbers attending). " (S13)* |
| | Facilitators & Barriers to Sustainability | *1. Barrier: "(..)it's almost like, well, you have innovation, but then you don't, there's no system, or not a very good system, for being able to convert that into something that actually works. That's what's missing." (P2); 2. Facilitator: "These groups are self-managed and self-directed, so we've kind of seeded these groups in different areas." (P6)* | *1 (Barrier) "We do not have funding for more resources." (S16)*<br><br>*2 (Facilitator) "We will still be able to build on the connections that we have made throughout the grant period such as with our new connections with the local autistic community." (S18)* |
| | Ongoing support needs | *"I think an opportunity to build more of a relationship with the funding team. And so, they could help us to understand where* | *"Longer-term funding commitments would help sustain the project- Short-term grants, while valuable, make it difficult to sustain* |

| | | | |
|---|---|---|---|
| | | *else we could go and use their expertise to signpost us to other funding, even if it was outside the government, would have been extremely useful." (P1)* | *and plan services for vulnerable groups. Multi-year funding would allow for stability, strategic growth, and better retention of staff and volunteers. [...] Offering transition or extension funding for projects awaiting renewal decisions could prevent service disruption for at-risk groups" (S52)* |
